# Pulmonary Artery Pressures and Mortality during VA ECMO: An ELSO Registry Analysis

**DOI:** 10.1101/2023.08.08.23293859

**Authors:** Clark G. Owyang, Brady Rippon, Felipe Teran, Daniel Brodie, Joaquin Araos, Daniel Burkhoff, Jiwon Kim, Joseph E. Tonna

**Affiliations:** Division of Pulmonary and Critical Care Medicine, Department of Medicine, NewYork-Presbyterian Hospital/Weill Cornell Medical College, New York, New York, USA; Department of Emergency Medicine, NewYork-Presbyterian Hospital/Weill Cornell Medical College, New York, New York, USA; Department of Population Health Sciences, Weill Cornell Medical College, New York, New York, USA; Department of Medicine, School of Medicine, Johns Hopkins University, Baltimore, MD, USA; Department of Clinical Sciences, College of Veterinary Medicine, Cornell University, Ithaca, NY, United States; Cardiovascular Research Foundation, New York, NY, USA; Division of Cardiology, Department of Medicine, Weill Cornell Medicine/New York Presbyterian Hospital, 525 East 68th Street, New York, NY, 10021, USA; Division of Cardiothoracic Surgery, Department of Surgery, University of Utah Health, Salt Lake City, UT, USA; Department of Emergency Medicine, University of Utah Health, Salt Lake City, UT, USA

**Keywords:** extracorporeal membrane oxygenation, extracorporeal life support, right ventricle, right ventricular dysfunction, right ventricular failure, right ventricle-pulmonary artery coupling

## Abstract

**Background:** Systemic hemodynamics and specific ventilator settings have been shown to predict survival during venoarterial extracorporeal membrane oxygenation (VA ECMO). While these factors are intertwined with right ventricular (RV) function, the independent relationship between RV function and survival during VA ECMO is unknown.

**Objectives:** To identify the relationship between RV function with mortality and duration of ECMO support.

**Methods:** Cardiac ECMO runs in adults from the Extracorporeal Life Support Organization (ELSO) Registry between 2010 and 2022 were queried. RV function was quantified via pulmonary artery pulse pressure (PAPP) for pre-ECMO and on-ECMO periods. A multivariable model was adjusted for Society for Cardiovascular Angiography and Interventions (SCAI) stage, age, gender, and concurrent clinical data (i.e., pulmonary vasodilators and systemic pulse pressure). The primary outcome was in-hospital mortality.

**Results:** A total of 4,442 ECMO runs met inclusion criteria and had documentation of hemodynamic and illness severity variables. The mortality rate was 55%; non-survivors were more likely to be older, have a worse SCAI stage, and have longer pre-ECMO endotracheal intubation times (P < 0.05 for all) than survivors. Improving PAPP from pre-ECMO to on-ECMO time (Δ PAPP) was associated with reduced mortality per 10 mm Hg increase (OR: 0.91 [95% CI: 0.86-0.96]; P=0.002). Increasing on-ECMO PAPP was associated with longer time on ECMO per 10 mm Hg (Beta: 15 [95% CI: 7.7-21]; P<0.001).

**Conclusions:** Early improvements in RV function from pre-ECMO values were associated with mortality reduction during cardiac ECMO. Incorporation of Δ PAPP into risk prediction models should be considered.

## Introduction/Background

Limited data exist on right heart function during cardiac support extracorporeal membrane oxygenation (ECMO).(1) Various critical care cohorts demonstrate that patients with right ventricular dysfunction (RVD) often have worse survival than those with isolated left ventricular (LV) dysfunction.(2–16) No study has examined pre- and on-ECMO right ventricular function (RV) or pulmonary artery (PA) pressures in relation to ECMO survival and duration of support. Whether RV function and associated PA pressures in cardiac support ECMO affect clinically important outcomes is unknown.

Recently, systemic pulse pressure and systolic blood pressure (i.e., left heart) improvements have been shown to predict improved survival in cardiac support ECMO (mostly venoarterial [VA] configuration) and ECMO after cardiac arrest.(17,18) Similar to systemic pulse pressure, pulmonary artery pulse pressure (PAPP [i.e., difference between pulmonary systolic and diastolic pressures]) is a clinically-relevant metric reflecting RV contractility. Generally, increasing PAPP (without a change in central venous pressure [CVP]) indicates greater forward flow. In contrast, low PAPP and high CVP predict RVD and failure in critically ill and mechanical circulatory support populations.(19–22) A recent iteration utilized the PAPP indexed to systolic pulmonary artery pressure (SPAP) with higher values predicting more favorable outcomes in pulmonary arterial hypertension cohorts.(23) The role of PAPP or PAPP indices as indicators of RV function remain undefined in ECMO.

It remains unknown if PAPP prognosticates survival, and if it should be a therapeutic target for patients during cardiac support ECMO. We hypothesized that early improvements in on-ECMO RV function, as quantified by increased PAPP and derived indices, would be associated with improved ECMO survival and with shorter ECMO run times. We queried the ELSO Registry, the largest global ECMO database, to investigate the role of RV function in cardiac support ECMO.

## Methods

Our analysis is reported according to the Strengthening the Reporting of Observational Studies in Epidemiology (STROBE) guidelines (**Supplemental eTable 1**).(24)

### Data Source

The ELSO Registry is a voluntary international registry with approximately 500 contributing centers across 60 countries and containing >200,000 patients over >30 years.(25,26) It is the largest international voluntary database of ECMO patients.(27) Standardized case report forms submitted by ELSO site data managers detail basic demographic data (age, weight, sex, race), pre- and on-ECMO ventilation data and circuit parameters as well as ICU location.(26) Real time data validation ensures data quality and accuracy of the Registry.(26,28) Data submission via ELSO site managers requires passage of the data entry exam with subsequent guidance via detailed instructions and standardized database definitions. Point-of-entry data assessment with error and validity checks along with full record validation at time of submission ensure completeness of mandatory fields. Internal validation has shown that only 1% of 190 reported fields were incorrect.(28,29) The Registry undergoes continuous multimodal auditing and has been previously validated.(28–30)

### Study Population

All adult (≥18 years) patients available from January 1, 2010 to May 1, 2022 who were supported with cardiac support ECMO were identified (**Supplemental eFigure 1**). Support type (i.e., cardiac, pulmonary, or extracorporeal cardiopulmonary resuscitation [ECPR]) does not necessarily correspond to the cannulation configuration (i.e., VV vs. VA); however, it describes the organ failure for which ECMO is initiated. Patients with pulmonary support-only types or ECPR and adult congenital heart disease (ACHD) patients were excluded. Cardiac support ECMO runs with missing or obviously inaccurate pulmonary artery (PA) pressures (e.g., where diastolic PA pressure is greater than systolic PA pressure) were excluded. ECMO runs with missing hemodynamic data, severity of illness data and demographics were also excluded.

To further define our cohort of interest for subgroup analysis, we identified the presence or absence of concomitant mechanical circulatory support (MCS) devices as well as cardiac surgery procedures based upon *Current Procedural Terminology* (CPT) codes. As previously described,(31) postoperative cardiac surgery patients were identified using CPT codes for heart or pericardium (33016–33999) excluding runs solely associated with codes for ECMO cannulation or daily management (33946–33989). Additionally, ventricular assist devices (VADs) and LV unloading CPT codes were identified as described in **Supplemental eTable 6**. Timing associated with MCS devices (i.e., LV unloading or intra-aortic balloon pumps) in relation to use specifically during concomitant ECMO run (rather than simply associated pre-cannulation or post-decannulation) was determined as previously described.(18,31) This retrospective analysis of the ELSO Registry was reviewed and approved by Weill Cornell Institutional Review Board.

### Hemodynamic Variables

Data on systemic blood pressure (systolic blood pressure [SBP], diastolic blood pressure [DBP], and pulse pressure [PP]) and pulmonary artery pressures (systolic pulmonary artery pressure [SPAP], diastolic pulmonary artery pressure [DPAP], and pulmonary artery pulse pressure [PAPP]) were collected at baseline pre-ECMO and at 24 hours post-ECMO initiation. Right atrial pressure (RAP [i.e., central venous pressure, CVP]) is not routinely collected within the ELSO Registry and thus was unavailable in our analysis.

### Exposure

Pulmonary artery pulse pressure (PAPP) was calculated as the difference between the SPAP and DPAP. Exposure variables reflecting RV function included the following: initial PAPP which is the pre-ECMO baseline; 24-hour on-ECMO PAPP which is an assessment closest to 24 hours after cannulation; and an interval change in PAPP (“Δ PAPP”) which is the difference between 24-hour on-ECMO value and pre-ECMO baseline. Consistent with standardized data collection of the ELSO Registry, pre-ECMO baseline variables were limited to no more than 6 hours prior to ECMO start time. On-ECMO variables were collected closest to 24 hours but no less than 18 hours and no more than 30 hours after start time. Missingness and handling of invalid data is detailed in the **Supplemental Methods** and **Supplemental eTable 2**.

### Outcomes

The primary outcome was in-hospital mortality for the index hospitalization. This was reflected on the ELSO Registry data form by discharged alive status as either “yes/no” or “transferred on ECMO.” Separately, the ECMO discontinuation reason is available on Registry forms with reasons including “poor prognosis followed by death” and “poor prognosis followed by unexpected survival”; however, this variable was not utilized in our binary primary outcome of survival or death.

Secondary outcome was duration of ECMO support calculated as the difference between the ECMO run’s date/time on and off.

### Covariates

For the main model, ELSO Registry variables determined to influence both PAPP and survival were included for analysis: age, gender, SCAI (Society for Cardiovascular Angiography) stage,(32) systemic pulse pressure and inhaled pulmonary vasodilators. Additional analysis included the main model with additional covariates of blood gases and mean airway pressure (**Supplemental eTable 3**). Time-specific variables (i.e., systemic pulse pressure, blood gases, and airway pressures) were collected in a similar manner as described previously for pre- vs. on-ECMO values (e.g., pre-ECMO PAPP effect on survival was evaluated with systemic pulse pressure covariate collected also during pre-ECMO interval). A modified SCAI stage classification was calculated as previously described.(32,33) Inhaled pulmonary vasodilator use is documented when the patient is on the medication for at least 6 hours within the 24-hour period prior to ECMO.

### Missing Data

Missing data is summarized in **Supplemental eTable 2**. ECMO run cases without complete documentation of PA pressures were excluded from analysis as detailed in **Supplemental eFigure 1**.

### Statistical Analysis

Baseline demographics, severity of illness, ECMO flow characteristics (4-hour and 24-hour), ECMO run times, and pre-ECMO intubation times were compared amongst survivors and non-survivors using the Pearson chi-square test for categorical variables (reported as proportions) as well as normally distributed continuous variables. Non-normally distributed continuous variables were reported as median (IQR) and were analyzed using the Wilcoxon rank sum test. Interval changes in blood pressure parameters were reported as integers (24-hour *minus* baseline) with respective negative or positive signs. For example, a systolic pressure of 70 mm Hg at 24 hours on ECMO from an initial pre-ECMO systolic blood pressure of 80 mm Hg was reported as a change of −10 mm Hg. Associations between baseline characteristics and the primary outcome variable were assessed using univariable logistic regression analyses. Multivariable logistic regression analysis including variables identified *a priori* to be related to right heart function and survival (age, race, SAVE score, ECMO flow at 24 hours, and blood pressure parameters) was performed similar to previously described work.(17) All p-values were calculated with respect to a two-sided alpha level of 0.05 which was deemed statistically significant. Statistical analysis was performed using the R statistical software package version 3.2.2 (R foundation, Vienna, Austria).

### Data Sharing

To facilitate research reproducibility, replicability, accuracy and transparency, the associated analytic code is available on the Open Science Foundation (OSF) repository, [DOI 10.17605/OSF.IO/82DKM] [https://osf.io/82dkm/]. The data that support the findings of this study are available from ELSO, upon request, to ELSO members; these data were used under license for the current study, and so are not publicly available. Data were de-identified in accordance with Section 164.514 of the Health Insurance Portability and Accountability Act (HIPAA).

## Results

### Study Population

Of the 37,147 eligible adult ECMO runs where the patients received cardiac support ECMO between 2010 and 2020 in the ELSO Registry and met our inclusion criteria, 4,997 runs contained valid measures for initial PAPP and 24-hour PAPP (i.e., complete documentation, non-zero and non-negative integers). An additional 555, runs were excluded for missing data (237 missing hemodynamic data; 211 missing SCAI stage; 104 missing age, and 3 missing ECMO time) leaving 4,442 ECMO runs for final analysis (**Table 1, Supplemental eFigure 1 and eTable 2**) in the main model.

**Table 1.** Patient Characteristics. Comparison of Baseline Characteristics Between Survivors and Non-survivors.

| Variable | N | Overall, N = 4,442<sup>1</sup> | Non-survivors, N = 2,459<sup>1</sup> | Survivors, N = 1,983<sup>1</sup> | P-value<sup>2</sup> |
| --- | --- | --- | --- | --- | --- |
| **Age, y** | 4,442 (100%) | 59 (49, 67) | 62 (53, 68) | 56 (44, 64) | **<0.001** |
| **Age group, y** | 4,442 (100%) |  |  |  |  |
| 18-38 |  | 562 (13%) | 198 (8.1%) | 364 (18%) | **<0.001** |
| 39-52 |  | 881 (20%) | 403 (16%) | 478 (24%) | **<0.001** |
| 53-62 |  | 1,291 (29%) | 721 (29%) | 570 (29%) | 0.7 |
| ≥63 |  | 1,708 (38%) | 1,137 (46%) | 571 (29%) | **<0.001** |
| **Women, %** | 4,442 (100%) | 1,379 (31%) | 763 (31%) | 616 (31%) | >0.9 |
| **Race** | 4,442 (100%) |  |  |  |  |
| White |  | 2,904 (65%) | 1,655 (67%) | 1,249 (63%) | **0.003** |
| Black |  | 541 (12%) | 271 (11%) | 270 (14%) | **0.009** |
| Hispanic |  | 301 (6.8%) | 153 (6.2%) | 148 (7.5%) | 0.10 |
| Asian |  | 207 (4.7%) | 98 (4.0%) | 109 (5.5%) | **0.018** |
| Other |  | 323 (7.3%) | 179 (7.3%) | 144 (7.3%) | >0.9 |
| Unknown |  | 166 (3.7%) | 103 (4.2%) | 63 (3.2%) | **0.077** |
| **Weight, kg** | 4,411 (99%) |  |  |  |  |
| ≤65 |  | 600 (14%) | 303 (12%) | 297 (15%) | **0.010** |
| 65-88 |  | 1,763 (40%) | 958 (39%) | 805 (41%) | 0.3 |
| ≥89 |  | 2,048 (46%) | 1,182 (48%) | 866 (44%) | **0.004** |
| **SCAI Stage** | 4,442 (100%) |  |  |  |  |
| B |  | 150 (3.4%) | 74 (3.0%) | 76 (3.8%) | 0.13 |
| C |  | 550 (12%) | 268 (11%) | 282 (14%) | **<0.001** |
| D |  | 3,555 (80%) | 1,995 (81%) | 1,560 (79%) | **0.041** |
| E |  | 187 (4.2%) | 122 (5.0%) | 65 (3.3%) | **0.005** |
| **Pre-ECLS Intubation, h** | 3,911 (88%) |  |  |  |  |
| ≤10 |  | 1,820 (47%) | 930 (43%) | 890 (52%) | **<0.001** |
| 11-29 |  | 1,054 (27%) | 596 (27%) | 458 (27%) | 0.6 |
| ≥30 |  | 1,037 (27%) | 662 (30%) | 375 (22%) | **<0.001** |
| **ELCS Characteristics** |  |  |  |  |  |
| Run-time, h | 4,442 (100%) | 136 (79, 222) | 144 (78, 244) | 124 (81, 199) | **<0.001** |
| Flow at 4h | 4,312 (97%) | 4.08 (3.50, 4.60) | 4.09 (3.50, 4.62) | 4.06 (3.50, 4.60) | 0.13 |
| Flow at 24h | 4,276 (96%) | 4.12 (3.57, 4.71) | 4.19 (3.62, 4.77) | 4.09 (3.50, 4.67) | **<0.001** |

<sup>1</sup>Median (IQR); n (%)<sup>2</sup>Wilcoxon rank sum test; Pearson's Chi-squared test
SCAI Stage = Society for Cardiovascular Angiography cardiogenic shock severity stage; ECLS = extracorporeal life support

The final study population had a median age of 59.1 years (IQR: 49-67 years) and 31% were female. The SCAI stages (as previously described(32)) distribution was as follows: stage B: 3.4%; stage C: 12%; stage D: 80%, and stage E: 4.2%. Time on mechanical ventilation prior to ECMO start time ranged from ≤ 10 hours (47% of our final cohort) to 11-29 hours (27%) and ≥ 30 hours (27%). ECMO run median time was 136 hours (IQR: 79-222 hours) with 144 hours (IQR: 78-244 hours) in non-survivors and 124 hours (81-199 hours) in survivors. Mortality in our final analytic cohort was 55%.

### Pre-ECMO, On-ECMO and Interval Change in Hemodynamics

**Table 2** shows the distribution of systemic and right heart hemodynamics separated into non-survivors and survivors for pre-ECMO, 24 hours on-ECMO, and interval change from pre- to on-ECMO values (Δ PAPP). The following trends in **Table 2** were observed prior to analysis with our multivariate model in **Table 3**.

**Table 2.**
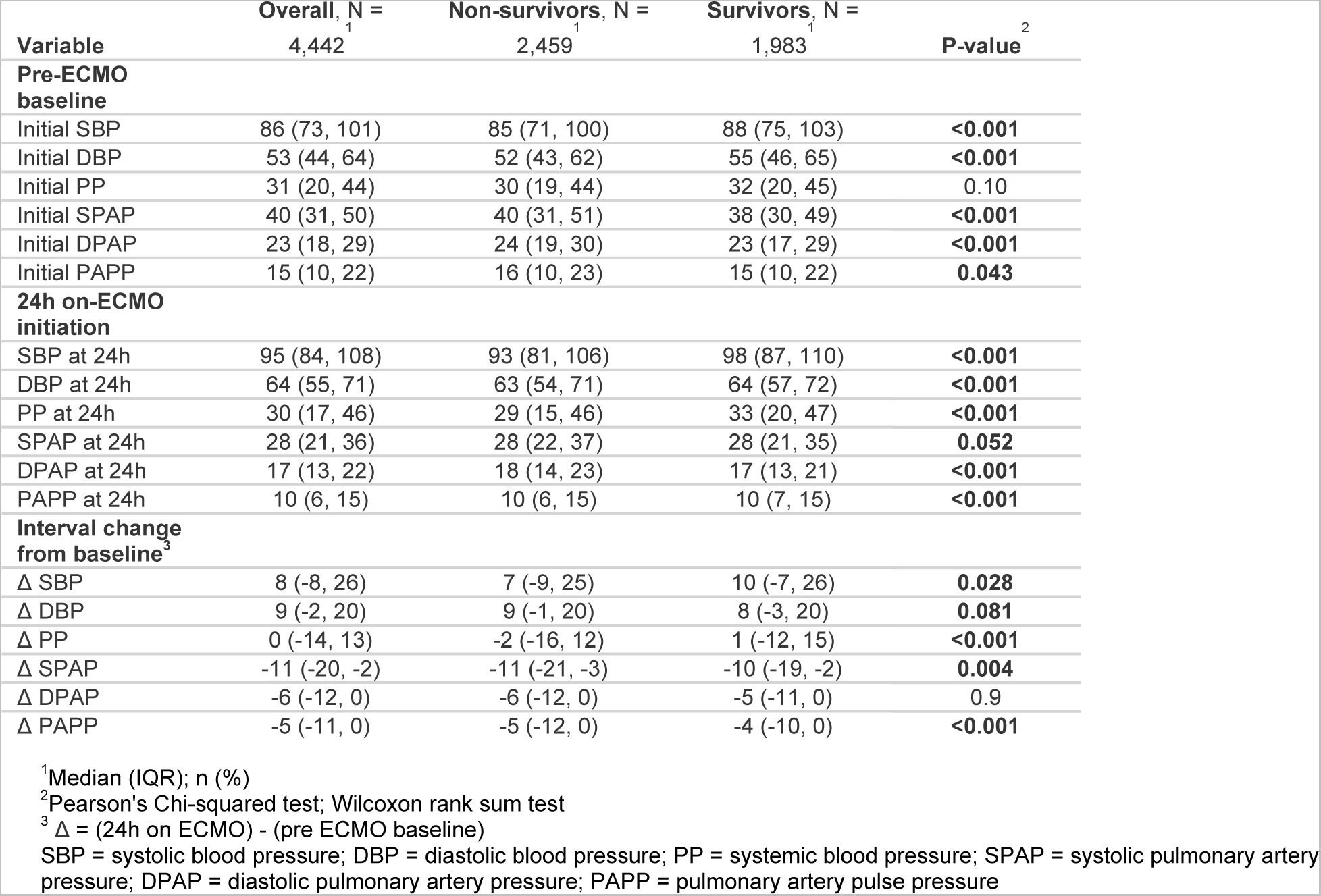
Hemodynamic Measurements: Non-survivors and Survivors.

**Table 3.** Right Heart Predictors of Mortality.

|  | Pre-ECMO PAPP |  |  | On-ECMO PAPP at 24h |  |  | Δ PAPP<sup>3</sup> |  |  |
| --- | --- | --- | --- | --- | --- | --- | --- | --- | --- |
|  | OR<sup>1</sup> | 95% CI | P-value | OR<sup>1</sup> | 95% CI | P-value | OR<sup>1</sup> | 95% CI | P-value |
| **PAPP** | 1.08<sup>2</sup> | 1.02, 1.14 | **0.006** | 0.98<sup>2</sup> | 0.90, 1.06 | 0.54 | 0.91<sup>2</sup> | 0.86, 0.96 | **0.002** |
| **PAPP quartiles<sup>4</sup>** |  |  |  |  |  |  |  |  |  |
| 1 | — | — |  | — | — |  | — | — |  |
| 2 | 0.97 | 0.81, 1.15 | 0.72 | 0.77 | 0.64, 0.92 | **0.005** | 0.87 | 0.73, 1.04 | 0.13 |
| 3 | 1.02 | 0.85, 1.22 | 0.83 | 0.75 | 0.63, 0.90 | **0.002** | 0.72 | 0.60, 0.85 | **<0.001** |
| 4 | 1.15 | 0.96, 1.37 | 0.14 | 0.79 | 0.66, 0.95 | **0.010** | 0.84 | 0.70, 0.99 | **0.044** |

<sup>1</sup>Adjusted for age, sex, SCAI stage, systemic pulse pressure, and inhaled pulmonary vasodilators
<sup>2</sup>Odds ratio per 10 mm Hg increase in PAPP
<sup>3</sup>Δ PAPP = difference between on-ECMO and pre-ECMO PAPP
<sup>4</sup>PAPP quartile (Q) ranges from [1, 10] for Q1, [10, 15] for Q2, [15, 22] for Q3, and [22, 129] for Q4; 24h PAPP ranges from [1, 6] for Q1, [6, 9] for Q2, [9, 14] for Q3, and [14, 110] for Q4.
PAPP = pulmonary artery pulse pressure

The pre-ECMO baseline median SBP (85 mm Hg vs. 88 mm Hg) and DBP (52 mm Hg vs. 55 mm Hg) were significantly lower among non-survivors compared with survivors (P < 0.001), whereas the PP was not (32 mm Hg vs. 30 mm Hg; P=0.10 [**Table 2**]). In contrast, pre-ECMO SPAP (40 mm Hg vs. 38 mm Hg), DPAP (24 mm Hg vs. 23 mm Hg), and PAPP (16 mm Hg vs. 15 mm Hg) were higher amongst non-survivors compared to survivors (P < 0.05 for all) prior to any analysis in our multivariate model.

Trends for 24 hours on-ECMO and interval change from pre- to on-ECMO time points are summarized with systemic pressures increasing with pulmonary artery pressures decreasing. At the time point of 24-hours on-ECMO (**Table 2**), all patients (whether non-survivor or survivor) had higher median SBP, DBP, and PP compared to all patients pre-ECMO baseline. With ECMO initiation, the right heart pressures (SPAP, DPAP, and PAPP) were correspondingly lower at 24-hours on ECMO compared to pre-ECMO baseline.

For interval change from pre- to on-ECMO, the systemic blood pressures trended upwards following ECMO; in contrast, the right heart pressures (SPAP, DPAP, and PAPP) generally trended downwards across non-survivors and survivors.

### Outcomes: Mortality & ECMO Run Time

Amongst the final cohort of 4,442 ECMO runs in our analysis, 55% of the cohort died during the index ECMO run. Mortality risk in the multivariate model is depicted as a function of PAPP at pre-ECMO, on-ECMO and Δ PAPP (difference between on-ECMO and pre-ECMO baseline) time points in **Table 3** and **Figure 1**.

**Figure 1.**
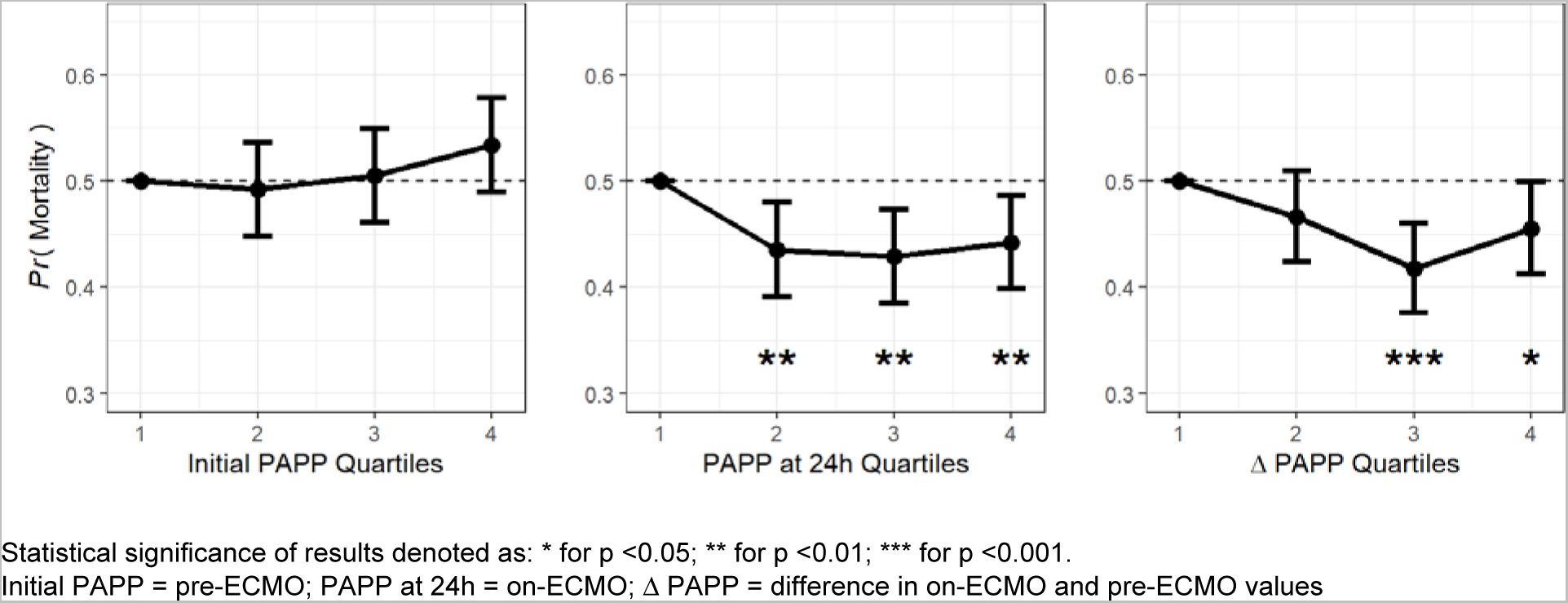
Adjusted Probability of Mortality for Pre-ECMO PAPP (box 1), On-ECMO PAPP (box 2) and Δ PAPP (box 3) by quartile.

On-ECMO PAPP and Δ PAPP were associated with improved odds of survival after correcting for demographics, severity of illness, systemic pulse pressure and inhaled pulmonary vasodilators (**Table 3**). Specifically, increasing quartiles of on-ECMO PAPP were associated with reduced odds of mortality compared to the lowest quartile (OR: 0.77 [95% CI: 0.64-0.92]; P=0.005, OR: 0.75 [95% CI: 0.63-0.90]; P=0.002, and OR: 0.79 [95% CI: 0.66-0.95]; P=0.010, respectively). Additionally, every 10 mm Hg increase in the Δ PAPP was associated with an odds ratio for in-hospital mortality of 0.91 (95% CI: 0.86-0.96; P=0.002). Reduced odds of mortality were similarly seen with every 10 mm Hg increase in Δ PAPP (OR: 0.89 [95% CI: 0.83-0.96]; P=0.002) in the supplemental model which augmented our main model via additional adjustment for blood gas values and mean airway pressure (**Supplemental eTable 3**). In contrast, higher pre-ECMO PAPP was associated with greater mortality for every 10 mm Hg increase (OR: 1.08 [95% CI: 1.02-1.14]; P=0.006). This was observed in both the main model (**Table 3**) as well as the supplemental model (**Supplemental eTable 3**).

Higher pre-ECMO and on-ECMO PAPP were associated with longer ECMO run time while Δ PAPP was not significantly associated with ECMO time (**Supplemental eTable 4**). Additionally, the direction of this relationship of higher PAPP and longer ECMO time held in preliminary analysis after removing patients who died on ECMO prior to adjustment for covariates.

### Subgroup Analysis: Cardiac Surgical Procedures, Ventricular Assist Device and LV Unloading

When our main multivariate model was applied to the subgroup of post-operative cardiac surgery patients (n = 1,414), on-ECMO PAPP at 24 hours associated with reduced mortality (**Supplemental eTable 5**) when assessed in 10 mm Hg increments (OR: 0.81 [95% CI: 0.69-0.93]; P=0.004). Cardiac surgery patients had lower mortality at 24 hours on-ECMO for the 3rd and 4th highest on-ECMO PAPP quartiles when compared to the lowest quartile (OR: 0.58 [95% CI: 0.42-0.79]; P<0.001 and OR: 0.73 [95% CI: 0.54-0.99]; P=0.04, respectively). PAPP improvements (“Δ PAPP”) per 10 mm Hg increments did not reach statistical significance (OR: 0.98 [95% CI: 0.88-1.09]; P=0.75 and OR: 0.97 [95% CI: 0.86-1.09]; P=0.61, respectively). The subgroup analysis within VAD or LV unloading populations showed no correlation between right heart function and survival (**Supplemental eTable 5**).

## Discussion

Our work shows that early on-ECMO surrogates of right heart function associate with improved survival in VA ECMO populations. After correcting for demographics; severity of illness; systemic pulse pressure; and inhaled pulmonary vasodilators, early pulmonary artery pressure metrics are independently associated with survival. On-ECMO PAPP (i.e., at 24-hours of ECMO time) and Δ PAPP (i.e., interval change on-ECMO from pre-ECMO value) were associated with statistically significant mortality reductions. In contrast, pre-ECMO PAPP was statistically associated with increased mortality. The risk of in-hospital mortality dropped incrementally with every 10 mm Hg increase in Δ PAPP once the patient was placed on ECMO. Given the distribution of on-ECMO PAPP across the dataset (i.e., the majority of patients decrease their PAPP from pre- to on-ECMO), these data may be more specifically interpreted as worsened mortality for those who fail to increase their on-ECMO PAPP. As most patients do not increase their PAPP once ECMO is initiated, our results can be interpreted as worse mortality for every 10 mmHg decrease in on-ECMO PAPP.

When analyzed by quartile, higher quartiles of both on-ECMO PAPP and Δ PAPP had lower odds of mortality when compared to the lowest quartile of each group, respectively. Further analysis showed that the incremental survival benefit for Δ PAPP was maintained with similar effect and with statistical significance after subsequent additional adjustment for blood gas variables and mean airway pressure in our supplemental model. To our knowledge, this analysis is the first to show a clinically relevant surrogate of RV function has an independent role in survival for cardiac support (mostly venoarterial configuration) ECMO.

These findings show statistical significance and present the potential clinical significance of the RV in cardiac critical care cohorts and ECMO populations which has been increasing investigated in recent years.(2,12) Current research shows that RV dysfunction (RVD) increases with shock severity and that RVD portends worse outcomes compared to LV dysfunction.(12) Additionally, the RV is affected directly by the LV and systemic hemodynamics as well as mechanical ventilation.(34–36) Both systemic hemodynamics and modifiable ventilator settings have been shown to be associated with survival in VA ECMO.(17,37) Our study results are consistent with this prior work using early cardiopulmonary metrics in prognosticating ECMO survival and show the importance of PA pressures (surrogates of RV function) in predicting survival. Additionally, our results provide novel large dataset support for the biventricular and heart-lung interactions described in recent ECMO simulation studies, providing new insight into RV function’s role in ECMO.(34)

In addition to investigating how on-ECMO RV function affected survival, we attempted to evaluate if preserved pre-ECMO RV function going into the ECMO run predicted better outcomes. Where survival improved with increased PAPP post-ECMO cannulation, the opposite effect was observed for pre-ECMO PAPP. Higher PAPP before cannulation showed statistically reduced odds of survival (i.e., higher mortality) in both the main model and in the supplemental model. While we did not have CVP to improve the accuracy of PAPP in reflecting forward flow of the right heart, it’s possible that higher pulmonary vascular resistance (PVR) was the underlying driver of increased mortality. The baseline PAPP between groups showed a small difference (1 mm Hg) and may not be clinically significant, but the retained relationship in multivariate analysis could be explained by the association of higher PVR with mortality, as can happen with pulmonary congestion from heart failure or from hypoxemia. Indeed, our data demonstrate lower systemic pressure among non survivors, as could be seen with worsened heart failure. Further, it has already been demonstrated among patients managed on VA ECMO that pre-ECMO PaO_2_ is nearly 10 mm Hg worse among non-survivors.(37)

As data exist regarding the role of RV function in weaning from VA ECMO, we expected to find higher PAPP to associate with reduced time on ECMO; however, this was not seen in our analysis.(38–40) Higher pre-ECMO and on-ECMO PAPP showed statistically significant increases for ECMO time. This finding was counter to our initial hypothesis and could be a function of either institution-specific practices/protocolized nature of ECMO weaning or from the biventricular interactions in ECMO. Specifically, high PAPP paradoxically has been shown to distend and impair left ventricle (LV) function in VA ECMO.(34) It is plausible that robust right heart pulse pressure both pre-ECMO and on-ECMO could overload or distend the LV leading to longer ECMO time, though our analysis was not designed to fully evaluate this.(34)

Given the physiologic interaction of PAPP and LV distension,(34) we investigated whether LV unloading or ventricular assist device affected our findings of right heart function and survival. Interestingly, despite the on-ECMO PAPP and PAPP improvement correlations with survival in the main model, the subgroup analysis within VAD or LV unloading populations showed no correlation between right heart function and survival.

Cardiac support ECMO has expanded in recent years. While survival prediction scores such as the SAVE score have been published, subsequent validation studies have found survival underestimation in some cohorts, which could lead to underuse of a potentially effective therapy.(41) Further, the SAVE score utilizes pre-ECMO variables to predict survival. Despite survival prediction via pre-ECMO variables, there remain questions about how to optimally prognosticate and manage on-ECMO hemodynamics.(42) Much of the published literature to date relates to left heart function, LV distension and systemic variables as cardinal considerations in VA ECMO.(17,42,43) There is little known about if and to what extent RV function plays a role in cardiac support ECMO. Our study is among the first to show how on-ECMO surrogates of right heart function may affect survival. Similar to work by Rali et al., the use of early on-ECMO hemodynamics could be used clinically to appropriately allocate resources or target individualized therapy. Future studies should provide comprehensive assessment of RV function via advanced echocardiography and/or pressure-volume loops. This will identify the true link between RV function and PAPP, so the latter can be implemented as a valid surrogate with prognostic value in this cohort.

## Study Limitations

While this analysis is among the first to evaluate right heart function in a large ECMO database, a large proportion of the initial cardiac support cohort did not contain pulmonary artery catheter data. From our initial raw data of 37,668 ECMO runs, PAPP for pre-ECMO and on-ECMO time periods was available in only 3% of cases. Previously reported analyses have focused on the left heart for analysis; similar to Rali et al., our analysis found early on-ECMO hemodynamics to statistically affect mortality.(17) Despite cohort limitations of the dataset, our study addresses an important gap in the literature.

Additionally, our choice for PAPP as exposure variable was based in the best-available data from ELSO Registry approximating the prognostic utility of the pulmonary artery pulsatility index (PAPi) seen in multiple populations.(44,45) However, we acknowledge that the PAPP is a surrogate measure of right heart function with less well-defined ability to predict mortality in other settings as more data are published on PAPi. Further, it is unknown whether PAPP is a sensitive/specific marker of RV function. However, we used these data as it was the closest related to RV function available from the Registry. Despite the unclear trend with mortality seen in the Blyth et al. analysis, the PAPP is the closest available variable in the Registry that approximates PAPi.(46) CVP as a variable is not collected in the ELSO Registry.

As the left and right heart interact, the lack of pulmonary capillary wedge pressure measurements in our dataset limits the ability to systematically assess the contribution of elevated left heart filling pressures on outcomes. Previous work has shown the relationship between higher PA pulse pressure (i.e. increasing right heart function or end-systolic elastance) and increasing LV end-diastolic pressures.(34) Approximately 88% of our cohort with available PA pulse pressure did not have concomitant PCWP metrics. Even more specifically, this analysis was unable to make the distinction between disease states with both elevated pulmonary capillary wedge pressure and diastolic pulmonary artery pressure versus disease states with uncoupling of the wedge and DPAP. Taken together, it is impossible to say what contribution left heart function or pre-existing pulmonary hypertension had on our findings. We acknowledge that without CVP or PCWP, the conclusions are not optimally specific in determining causality specific to right heart function and mortality.

Further missingness was experienced in LV unloading devices subgroup via the documentation method for the device. CPT codes for various types of LV unloading devices were queried in a manner to be specific about their use prior to ECMO decannulation to avoid capturing LV unloading (i.e. percutaneous axial pump) devices post-ECMO. LV unloading devices certainly could have further influenced our findings as they influence left heart distension and subsequent right heart and lung parenchyma physiology. Our supplemental model incorporated blood gas data and mean airway pressure and still found similar on-ECMO right heart effects.

Finally, we also did not include adverse effects of ECMO (e.g., major bleeding, stroke, or circuit-related complications). As previously mentioned, our dataset was limited to the fraction of the entire cohort who had pulmonary artery catheter data. As this analysis is novel in its examination of right heart variables on ECMO, we chose to limit our work to outcomes of mortality and time on ECMO.

Despite the limitations, the central findings of our analysis hold in both the main model and our supplemental analysis. For the main model, 51% of our cohort was missing data for mean airway pressure, prompting our supplemental analysis. Our supplemental analysis was performed on the ~ 3,000 ECMO runs who had main analysis covariates as well as the blood gas and MAP covariates available. Our main model and supplemental model show similar incremental mortality reduction by PAPP improvements from pre- to on-ECMO status.

## Conclusion

Early improvements in PAPP from pre-ECMO values were associated with statistically significant mortality reduction during cardiac ECMO. Correspondingly and more specifically, failure to improve PAPP upon ECMO initiation is associated with worsened mortality.

Incorporation of Δ PAPP into risk prediction models for cardiac ECMO should be further investigated and considered.

## Clinical Perspectives

Clinical Implications: Surrogates of right heart function should be considered for incorporation into risk prediction models for cardiac support ECMO.

Competency in Medical Knowledge: Although prognostication scores have incorporated systemic hemodynamics and ventilator settings for ECMO risk stratification, there are limited data on the role of the right heart for early post-cannulation prognostication.

Translational Outlook: Despite its physiologic relevance, right heart variables have not been included in the analysis of cardiac support ECMO patients. Right heart function is increasingly identified in critically ill patients to be prognostically important, but further research is required to allow clinicians to better guide ECMO therapy.

## Abbreviations

ECMO: extracorporeal membrane oxygenation
ELSO: Extracorporeal Life Support Organization
ECPR: extracorporeal cardiopulmonary resuscitation
LV: left ventricle
PAPP: pulmonary artery pulse pressure
SPAP: systolic pulmonary artery pressure
DPAP: diastolic pulmonary artery pressure
PA: pulmonary artery
PVR: pulmonary vascular resistance
CVP: central venous pressure
RV: right ventricle
RVD: right ventricular dysfunction
VV: venovenous
VA: venoarterial
CPT: Current Procedural Terminology
ICD-9/10: International Classification of Diseases, Ninth Revision/Tenth Revision
MAP: mean airway pressure
ACHD: adult congenital heart disease
MCS: mechanical circulatory support
VAD: ventricular assist device
SCAI: Society for Cardiovascular Angiography

## Notes

<u>Conflicts of Interest and Funding Sources:</u> Dr. Tonna is supported by a Career Development Award from the National Institutes of Health/National Heart, Lung, And Blood Institute (K23 HL141596). Dr. Tonna is the Chair-elect of the Registry Committee of the Extracorporeal Life Support Organization (ELSO). Dr. Brodie receives research support from and consults for LivaNova. He has been on the medical advisory boards for Abiomed, Xenios, Medtronic, Inspira and Cellenkos. He is the President-elect of ELSO, the Chair of the Executive Committee of the International ECMO Network (ECMONet), and he writes for UpToDate. None of the funding sources were involved in the design or conduct of the study, collection, management, analysis or interpretation of the data, or preparation, review or approval of the manuscript. No conflicts of interest reported.

### Competing Interest Statement

The authors have declared no competing interest.

### Funding Statement

Dr. Tonna is supported by a Career Development Award from the National Institutes of Health/National Heart, Lung, And Blood Institute (K23 HL141596). Dr. Tonna is the Chair-elect of the Registry Committee of the Extracorporeal Life Support Organization (ELSO). Dr. Brodie receives research support from and consults for LivaNova. He has been on the medical advisory boards for Abiomed, Xenios, Medtronic, Inspira and Cellenkos. He is the President-elect of ELSO, the Chair of the Executive Committee of the International ECMO Network (ECMONet), and he writes for UpToDate. None of the funding sources were involved in the design or conduct of the study, collection, management, analysis or interpretation of the data, or preparation, review or approval of the manuscript. No conflicts of interest reported.

### Author Declarations

This retrospective analysis of the ELSO Registry was reviewed and approved by Weill Cornell Institutional Review Board.

## References

1. Kuroda T, Miyagi C, Fukamachi K, Karimov JH. Mechanical circulatory support devices and treatment strategies for right heart failure. Front Cardiovasc Med 2022;9:951234.

2. Sato R, Dugar S, Cheungpasitporn W et al. The impact of right ventricular injury on the mortality in patients with acute respiratory distress syndrome: a systematic review and meta-analysis. Crit Care 2021;25:172.

3. Kanwar MK, Everett KD, Gulati G, Brener MI, Kapur NK. Epidemiology and management of right ventricular-predominant heart failure and shock in the cardiac intensive care unit. Eur Heart J Acute Cardiovasc Care 2022;11:584–594.

4. Ramjee V, Grossestreuer AV, Yao Y et al. Right ventricular dysfunction after resuscitation predicts poor outcomes in cardiac arrest patients independent of left ventricular function. Resuscitation 2015;96:186–91.

5. Tabi M, Burstein BJ, Anavekar NS, Kashani KB, Jentzer JC. Associations of Vasopressor Requirements With Echocardiographic Parameters After Out-of-Hospital Cardiac Arrest. J Intensive Care Med 2022;37:518–527.

6. Tabi M, Singam NSV, Wiley B, Anavekar N, Barsness G, Jentzer JC. Echocardiographic Characteristics of Cardiogenic Shock Patients with and Without Cardiac Arrest. J Intensive Care Med 2022:8850666221105236.

7. Jentzer JC, Wiley BM, Reddy YNV, Barnett C, Borlaug BA, Solomon MA. Epidemiology and outcomes of pulmonary hypertension in the cardiac intensive care unit. Eur Heart J Acute Cardiovasc Care 2022;11:230–241.

8. Corica B, Marra AM, Basili S et al. Prevalence of right ventricular dysfunction and impact on all-cause death in hospitalized patients with COVID-19: a systematic review and meta-analysis. Sci Rep 2021;11:17774.

9. Jentzer JC, Anavekar NS, Reddy YNV et al. Right Ventricular Pulmonary Artery Coupling and Mortality in Cardiac Intensive Care Unit Patients. J Am Heart Assoc 2021;10:e019015.

10. Li Y, Li H, Zhu S et al. Prognostic Value of Right Ventricular Longitudinal Strain in Patients With COVID-19. JACC Cardiovasc Imaging 2020;13:2287–2299.

11. Dupont M, Mullens W, Skouri HN et al. Prognostic role of pulmonary arterial capacitance in advanced heart failure. Circ Heart Fail 2012;5:778–85.

12. Burstein B, van Diepen S, Wiley BM, Anavekar NS, Jentzer JC. Biventricular Function and Shock Severity Predict Mortality in Cardiac ICU Patients. Chest 2022;161:697–709.

13. Naksuk N, Tan N, Padmanabhan D et al. Right Ventricular Dysfunction and Long-Term Risk of Sudden Cardiac Death in Patients With and Without Severe Left Ventricular Dysfunction. Circ Arrhythm Electrophysiol 2018;11:e006091.

14. Jain P, Thayer KL, Abraham J et al. Right Ventricular Dysfunction Is Common and Identifies Patients at Risk of Dying in Cardiogenic Shock. J Card Fail 2021;27:1061–1072.

15. Lanspa MJ, Cirulis MM, Wiley BM et al. Right Ventricular Dysfunction in Early Sepsis and Septic Shock. Chest 2021;159:1055–1063.

16. Homan EA, Devereux RB, Tak KA et al. Impact of acute TTE-evidenced cardiac dysfunction on in-hospital and outpatient mortality: A multicenter NYC COVID-19 registry study. PLoS One 2023;18:e0283708.

17. Rali AS, Ranka S, Butcher A et al. Early Blood Pressure Variables Associated With Improved Outcomes in VA-ECLS: The ELSO Registry Analysis. JACC Heart Fail 2022;10:397–403.

18. Tonna JE, Selzman CH, Bartos JA et al. The Association of Modifiable Postresuscitation Management and Annual Case Volume With Survival After Extracorporeal Cardiopulmonary Resuscitation. Crit Care Explor 2022;4:e0733.

19. Kang G, Ha R, Banerjee D. Pulmonary artery pulsatility index predicts right ventricular failure after left ventricular assist device implantation. J Heart Lung Transplant 2016;35:67–73.

20. Kane CJ, Salama AA, Pislaru C, Kane GC, Pislaru SV, Lin G. Low Pulmonary Artery Pulsatility Index by Echocardiography Is Associated With Increased Mortality in Pulmonary Hypertension. J Am Soc Echocardiogr 2023;36:189–195.

21. Mazimba S, Welch TS, Mwansa H et al. Haemodynamically Derived Pulmonary Artery Pulsatility Index Predicts Mortality in Pulmonary Arterial Hypertension. Heart Lung Circ 2019;28:752–760.

22. Zern EK, Wang D, Rambarat P et al. Association of Pulmonary Artery Pulsatility Index With Adverse Cardiovascular Events Across a Hospital-Based Sample. Circ Heart Fail 2022;15:e009085.

23. Mwansa H, Bilchick KC, Parker AM et al. Decreased pulmonary arterial proportional pulse pressure is associated with increased mortality in group 1 pulmonary hypertension. Clin Cardiol 2017;40:988–992.

24. von Elm E, Altman DG, Egger M et al. Strengthening the Reporting of Observational Studies in Epidemiology (STROBE) statement: guidelines for reporting observational studies. BMJ 2007;335:806–8.

25. Nasr VG, Raman L, Barbaro RP et al. Highlights from the Extracorporeal Life Support Organization Registry: 2006-2017. ASAIO J 2019;65:537–544.

26. ELSO. Resources for Submission of Patient Data to the ELSO Registry. ELSO, 2023:Registry Database Definitions and Forms.

27. Nakashima T, Ogata S, Noguchi T et al. Association of intentional cooling, achieved temperature and hypothermia duration with in-hospital mortality in patients treated with extracorporeal cardiopulmonary resuscitation: An analysis of the ELSO registry. Resuscitation 2022;177:43–51.

28. Lorusso R, Alexander P, Rycus P, Barbaro R. The Extracorporeal Life Support Organization Registry: update and perspectives. Ann Cardiothorac Surg 2019;8:93–98.

29. Dalton HJ, Butt WW. Extracorporeal life support: an update of Rogers’ Textbook of Pediatric Intensive Care. Pediatr Crit Care Med 2012;13:461–71.

30. Schmidt M, Bailey M, Sheldrake J et al. Predicting survival after extracorporeal membrane oxygenation for severe acute respiratory failure. The Respiratory Extracorporeal Membrane Oxygenation Survival Prediction (RESP) score. Am J Respir Crit Care Med 2014;189:1374–82.

31. Owyang CG, Donnat C, Brodie D et al. Similarities in extracorporeal membrane oxygenation management across intensive care unit types in the United States: An analysis of the Extracorporeal Life Support Organization Registry. Artif Organs 2022.

32. Jentzer JC, Baran DA, Bohman JK et al. Cardiogenic shock severity and mortality in patients receiving venoarterial extracorporeal membrane oxygenator support. Eur Heart J Acute Cardiovasc Care 2022.

33. Thayer KL, Zweck E, Ayouty M et al. Invasive Hemodynamic Assessment and Classification of In-Hospital Mortality Risk Among Patients With Cardiogenic Shock. Circ Heart Fail 2020;13:e007099.

34. Donker DW, Sallisalmi M, Broome M. Right-Left Ventricular Interaction in Left-Sided Heart Failure With and Without Venoarterial Extracorporeal Membrane Oxygenation Support-A Simulation Study. ASAIO J 2021;67:297–305.

35. Magder S, Slobod D, Assanangkornchai N. Right Ventricular Limitation: A Tale of Two Elastances. Am J Respir Crit Care Med 2023;207:678–692.

36. Slobod D, Assanangkornchai N, Magder S. Comparison of Right Ventricular Loading by Lung Inflation During Passive and Assisted Mechanical Ventilation. Am J Respir Crit Care Med 2023.

37. Tonna JE, Selzman CH, Bartos JA et al. The association of modifiable mechanical ventilation settings, blood gas changes and survival on extracorporeal membrane oxygenation for cardiac arrest. Resuscitation 2022;174:53–61.

38. Huang KC, Lin LY, Chen YS, Lai CH, Hwang JJ, Lin LC. Three-Dimensional Echocardiography-Derived Right Ventricular Ejection Fraction Correlates with Success of Decannulation and Prognosis in Patients Stabilized by Venoarterial Extracorporeal Life Support. J Am Soc Echocardiogr 2018;31:169–179.

39. Trahanas JM, Li SS, Crowley JC et al. How to Turn It Down: The Evidence and Opinions Behind Adult Venoarterial Extracorporeal Membrane Oxygenation Weaning. ASAIO J 2021;67:964–972.

40. Kim D, Park Y, Choi KH et al. Prognostic Implication of RV Coupling to Pulmonary Circulation for Successful Weaning From Extracorporeal Membrane Oxygenation. JACC Cardiovasc Imaging 2021;14:1523–1531.

41. Amin F, Lombardi J, Alhussein M et al. Predicting Survival After VA-ECMO for Refractory Cardiogenic Shock: Validating the SAVE Score. CJC Open 2021;3:71–81.

42. Tanaka D, Shimada S, Mullin M, Kreitler K, Cavarocchi N, Hirose H. What Is the Optimal Blood Pressure on Veno-Arterial Extracorporeal Membrane Oxygenation? Impact of Mean Arterial Pressure on Survival. ASAIO J 2019;65:336–341.

43. Rao P, Khalpey Z, Smith R, Burkhoff D, Kociol RD. Venoarterial Extracorporeal Membrane Oxygenation for Cardiogenic Shock and Cardiac Arrest. Circ Heart Fail 2018;11:e004905.

44. Cesini S, Bhagra S, Pettit SJ. Low Pulmonary Artery Pulsatility Index Is Associated With Adverse Outcomes in Ambulatory Patients With Advanced Heart Failure. Journal of Cardiac Failure 2020;26:352–359.

45. Korabathina R, Heffernan KS, Paruchuri V et al. The pulmonary artery pulsatility index identifies severe right ventricular dysfunction in acute inferior myocardial infarction. Catheter Cardiovasc Interv 2012;80:593–600.

46. Blyth KG, Syyed R, Chalmers J et al. Pulmonary arterial pulse pressure and mortality in pulmonary arterial hypertension. Respir Med 2007;101:2495–501.

